# A Comprehensive Epithelial Tubo-Ovarian Cancer Risk Prediction Model Incorporating Genetic and Epidemiological Risk Factors

**DOI:** 10.1101/2020.12.04.20244046

**Authors:** Andrew Lee, Xin Yang, Jonathan Tyrer, Aleksandra Gentry-Maharaj, Andy Ryan, Nasim Mavaddat, Alex P. Cunningham, Tim Carver, Stephanie Archer, Goska Leslie, Jatinderpal Kalsi, Faiza Gaba, Ranjit Manchanda, Simon A. Gayther, Susan J. Ramus, Fiona M. Walter, Marc Tischkowitz, Ian Jacobs, Usha Menon, Douglas F. Easton, Paul P.D. Pharoah, Antonis C. Antoniou

**Author notes:** **Corresponding author:** Antonis C. Antoniou, Centre for Cancer Genetic Epidemiology, University of Cambridge, Strangeways Research Laboratory, Worts Causeway, Cambridge, CB1 8RN, UK. joint first authors (Andrew Lee, Xin Yang).

## Abstract

**Background:** Epithelial tubo-ovarian cancer (EOC) has high mortality partly due to late diagnosis. Prevention is available but may be associated with adverse effects. A multifactorial risk model based on known genetic and epidemiological risk factors (RFs) for EOC can help identify females at higher risk who could benefit from targeted screening and prevention.

**Methods:** We developed a multifactorial EOC risk model for females of European ancestry incorporating the effects of pathogenic variants (PVs) in *BRCA1, BRCA2, RAD51C, RAD51D* and *BRIP1*, a polygenic risk score (PRS) of arbitrary size, the effects of RFs and explicit family history (FH) using a synthetic model approach. The PRS, PV and RFs were assumed to act multiplicatively.

**Results:** Based on a currently available PRS for EOC that explains 5% of the EOC polygenic variance, the estimated lifetime risks under the multifactorial model in the general population vary from 0.5% to 4.6% for the 1^st^ to 99^th^ percentiles of the EOC risk-distribution. The corresponding range for females with an affected first-degree relative is 1.9% to 10.3%. Based on the combined risk distribution, 33% of RAD51D PV carriers are expected to have a lifetime EOC risk of less than 10%. RFs provided the widest distribution, followed by the PRS. In an independent partial model validation, absolute and relative 5-year risks were well-calibrated in quintiles of predicted risk.

**Conclusion:** This multifactorial risk model can facilitate stratification, in particular among females with FH of cancer and/or moderate- and high-risk PVs. The model is available via the CanRisk Tool (www.canrisk.org).

## INTRODUCTION

Epithelial tubo-ovarian cancer (EOC), the seventh most common cancer in females globally, is often diagnosed at late stage and is associated with high mortality. There were 7,443 new cases of EOC and 4,116 deaths from EOC annually in the UK in 2015-2017.[1] Early detection could lead to an early-stage diagnosis, enabling curative treatment and reducing mortality. Annual multimodal screening using a longitudinal serum CA125 algorithm in females from the general population resulted in significantly more females diagnosed with early-stage disease but without a significant reduction in mortality.[2] Four-monthly screening using the same multimodal approach also resulted in a stage shift in females at high risk (>10% lifetime risk of EOC).[3] Currently, risk-reducing bilateral salpingo-oophorectomy (RRSO), upon completion of their families, remains the most effective prevention option,[4] and it has been recently suggested that RRSO would be cost-effective in postmenopausal females at >4% lifetime EOC risk.[5, 6] Beyond surgical risk, bilateral oophorectomy may be associated with increased cardiovascular mortality[7] and a potential increased risk of other morbidities such as parkinsonism, dementia, cardiovascular disease and osteoporosis,[8, 9] particularly in those who do not take HRT.[10] Therefore, it is important to target such prevention approaches to those at increased risk, who are most likely to benefit.

Over the last decade, there have been significant advances in our understanding of susceptibility to EOC. After age, family history (FH) is the most important risk factor for the disease. Approximately 35% of the observed familial relative risk (FRR) can be explained by rare pathogenic variants (PVs) in the *BRCA1, BRCA2, RAD51C, RAD51D* and *BRIP1* genes.[11-14] Recent evidence suggests that *PALB2, MLH1, MSH2* and *MSH6* are also involved in the EOC genetic susceptibility.[14-16] Common variants, each of small effect, identified through genome-wide association studies (GWAS),[17, 18] explain a further 4%. Several epidemiological risk factors (RFs) are also known to be associated with EOC risk, including use of menopausal hormone therapy (MHT), body mass index (BMI), history of endometriosis, use of oral contraception, tubal ligation and parity.[19-24] Despite these advances, those at high risk of developing EOC are currently identified mainly through FH of the disease or on the basis of having PVs in *BRCA1* and *BRCA2*. However, more specific risk prediction could be achieved by combining data on all known epidemiological and genetic risk factors. The published EOC prediction models consider either RFs[22, 23, 25] or common variants.[22, 26] No published EOC risk prediction model takes into account the simultaneous effects of the established EOC susceptibility genetic variants (rare and common), residual FH and other known RFs.

Using complex segregation analysis, we previously developed an EOC risk prediction algorithm that considered the effects of PVs in *BRCA1* and *BRCA2* and explicit FH of EOC and breast cancer (BC).[11] The algorithm modelled the residual, unexplained familial aggregation using a polygenic model that captured other unobserved genetic effects. The model did not explicitly include the effects of other established intermediate-risk PVs in genes such as *RAD51C, RAD51D* and *BRIP1*,[12-14, 27] which are now included on routine gene panel tests, the effects of recently developed EOC PRSs or the known RFs.

Here we present a methodological framework for extending this model to incorporate the explicit effects of PVs in *RAD51C, RAD51D* and BRIP1 for which reliable age-specific EOC risk estimates are currently available, up-to-date polygenic risk scores (PRS) and the known EOC RFs (Table 1). We used this multifactorial model to evaluate the impact of negative predictive testing in families with rare PVs and to assess the extent of EOC risk stratification that can be achieved in the general population, females with a FH of EOC and those carrying rare PVs. We evaluated the performance of a subset of this model in the UK Collaborative Trial of Ovarian Cancer Screening (UKCTOCS),[2] where females from the general population were followed up prospectively.

**Table 1.** Summary of components of the EOC risk model.

| Risk factor group | Risk factor category | Comments |
| --- | --- | --- |
| **Family history** | Explicit family history of ovarian and other cancers (breast, prostate, male breast, pancreatic). | Considers families of arbitrary size and structure, including affected and unaffected relatives. |
| **Sex** |  | Sex of all family members |
| **Age** |  | Ages at cancer diagnosis or current ages/age at death of family members. |
| **Genetic Factors** |  |  |
| Rare truncating/pathogenic variants | *BRCA1* |  |
|  | *BRCA2* |  |
|  | *RAD51D* |  |
|  | *RAD51C* |  |
|  | *BRIP1* |  |
| Common genetic variants | Polygenic risk score | Explaining 5.0% of the polygenic variance. |
| Unobserved genetic effects | Residual polygenic component | Accounts for the residual familial aggregation of EOC. |
| **Lifestyle/hormonal/reproductive** |  |  |
|  | Height | Measured in cm (5 categories) |
|  | Body Mass Index | Measured in kg/m<sup>2</sup> (3 categories) |
|  | Parity | Number of live births (3 categories) |
|  | Endometriosis | Yes/No |
|  | Use of oral contraception | Years of use (5 categories) |
|  | Use of Hormone Replacement Therapy | Never/Ever |
|  | Tubal Ligation | Yes/No |
| **Breast tumour pathology** | Oestrogen, Progesterone, HER2 Receptor, CK14, CK5/6 Status | As implemented in the BOADICEA breast cancer model |
| **Demographic factors** |  |  |
| Country of Origin | Country | Defines the underlying incidences used. |
| Birth Cohort | Defined by the person's year of birth | 8 calendar year-specific sets of incidences. |
| Family Ethnicity | Ashkenazi Jewish Origin |  |

## METHODS

### EOC risk prediction model development

No large datasets are currently available that include data on all known genetic and other EOC RFs. Therefore, we used a synthetic approach, described previously,[28] to extend our previous EOC model[11] by capitalising on published estimates of the associations of each RF with EOC. This approach was shown to provide valid risk estimates in the case of BC.[28-30]

Under the assumption that the effects of rare PVs, RFs and polygenic component are multiplicative on EOC risk, the incidence at age *t* for individual *i* was modelled as:

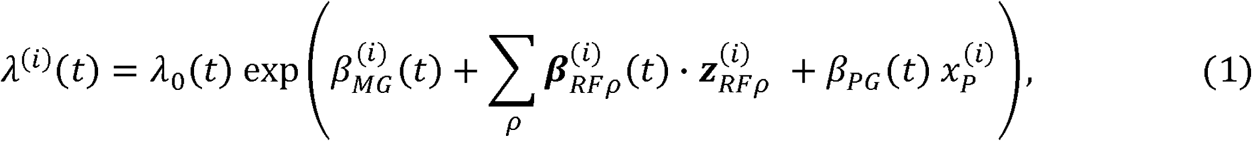

*λ*_0_(t) is the baseline incidence. 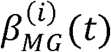 is the age-specific log-relative risk (log-RR) associated with individual *i*’s PV carrier status (explained below), relative to the baseline. The log-RR for non-carriers is 0. 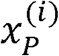 is the polygenotype for individual *i*, assumed to follow a standard normal distribution in the general population and *β*_*PG*_ (*t*) is the age-specific log-RR associated with the polygene, relative to the baseline incidence. 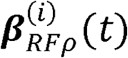 is the array of the log-RRs associated with risk-factor *ρ* at age *t*, which may depend on PV carrier status, and 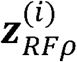 is the corresponding indicator array showing the category of risk-factor *ρ* for the individual. The baseline incidence was determined by constraining the overall incidences to agree with the population EOC incidence. To allow appropriately for missing RF information, only those RFs measured on a given individual are considered.

### Major gene effects

To include the effects of *RAD51D, RAD51C* and *BRIP1*, we used the approach described previously where PVs in these genes were assumed to be risk alleles of a single MG locus.[31] A dominant model of inheritance was assumed for all rare PVs. To define the penetrance, we assumed the following order of dominance when an individual carried more than one PV (that is, the risk was determined by the highest risk PV and any lower risk PVs ignored): *BRCA1, BRCA2, RAD51D, RAD51C, BRIP1*.[31] The population allele frequencies for RAD51D, RAD51C and BRIP1 and EOC RRs were obtained from published data (Table s3).[14, 27] Although PVs in *PALB2, MLH1, MSH2* and MSH6 have been reported to be associated with EOC risk, PVs in *MLH1, MHS2* and *MSH6* are primarily associated with risk of specific subtypes of EOC (endometrioid and clear cell),[15] and at the time of development, precise EOC age-specific risk estimates for *PALB2* PV carriers were not available. Therefore, these were not considered at this stage.

### Epidemiological Risk Factors

The RFs incorporated into the model include parity, use of oral contraception and MHT, endometriosis, tubal ligation, BMI and height. We assumed that the RFs were categorical and that an individual’s category was fixed for their lifetime, although the RRs were allowed to vary with age. The RR estimates used in equation (1) and population distributions for each RF were obtained from large-scale external studies and from national surveillance data sources, using a synthetic approach as previously described.[28] Where possible, we used RR estimates that were adjusted for the other RFs included in the model and distributions from the UK. Details of the population distributions and RRs used in the model are given in Table s2. As in BOADICEA,[28] in order to decrease the runtime, we combined the RFs with age-independent RRs into a single factor (specifically parity, tubal ligation, endometriosis, BMI and height).

### Incorporating Polygenic Risk Scores

We included an EOC susceptibility PRS, assumed to form part of the polygene, using the methods previously developed.[11, 28] The polygenic component decomposes into a measured component due to the PRS (*x*_*PRS*_) and an unmeasured component representing other familial effects (*x*_*R*_):

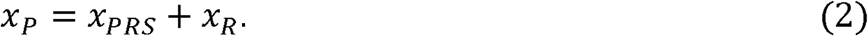

*x*_*PRS*_ summarises the effects of multiple common variants and is assumed normally distributed with mean 0 and variance *α*^2^ in the general population, with 0 ≤ *α* ≤ 1. The parameter *α*^2^ is the proportion of the overall polygenic variance (after excluding the effects of all MGs) explained by the PRS. *x*_*R*_ is normally distributed with mean 0 and variance 1− *α*^2^. The approach used to calculate *α*^2^ is described in the Supplementary Material. This implementation allows the effect size of the PRS to be dynamically varied, allowing an arbitrary PRS.

Here, for illustrating the model’s risk-stratification potential, we considered the latest validated EOC PRS developed by the Ovarian Cancer Association Consortium,[32] which is composed of 36 variants (Table s1) and has a log-variance of 0.099, accounting for 5.0% of the overall polygenic variance in the model. This 36 variant PRS was found to perform equally well as those comprised of more variants based on penalised regression or Bayesian approaches.[32]

### Other model components

The previous version[11] modelled the incidence of EOC and first female BC. To align with BOADICEA,[28] the model was extended to take account of female contralateral BC and the associations of *BRCA1/2* PVs with pancreatic cancer, male BC and prostate (Supplementary Methods).

### Model Validation

#### Study subjects

A partial model validation was carried out in a nested case-control sample of females of self-reported European ancestry participating in UKCTOCS. Based on the data available, we were able to validate the model on the basis of FH, PRS and RFs. Details of the UKCTOCS study design, blood sampling process, DNA extraction and processing, variant selection, genotyping and data processing are described in the Supplementary Methods and published elsewhere.[33] The study was approved by local ethical review committees. In summary, the following self-reported information was collected at recruitment and used for model validation: parity, use of oral contraception and MHT, tubal ligation, BMI and height (Table s4). As the study participants were genotyped for only 15 SNPs known at the time to be associated with EOC risk, it was not possible to use the more recently developed PRS for model validation. Instead, as the model can accommodate an arbitrary PRS, a PRS based on the on the 15 available SNPs was used[33] (Table s5), for which *α*^2^ = 0.037. The UKCTOCSS study participants were independent of the sets used to generate this PRS.[33] Study participants were not screened for PVs in *BRCA1, BRCA2, RAD51C, RAD51D* or *BRIP1*.

#### Pedigree construction

The UKCTOCS recruitment questionnaire collected only summary data on FH of BC and EOC. Since the risk algorithm uses explicit FH information, these data were used to reconstruct the pedigrees, which included information on incidences in the first- and second-degree relatives (Supplementary Methods).

#### Statistical analysis

All UKCTOCS participants were followed using electronic health record linkage to national cancer and death registries. For this study, they were censored at either: their age at EOC, their age at other (non-EOC) first cancer diagnosis, their age at death or age 79. To assess the model performance, a weighted approach was used whereby each participant was assigned a sampling weight based on the inverse of the probability of being included in the nested case-control study, given their disease status. Since all incident cancer cases were included, cases were assigned a weight of 1. The cases were matched to two random controls (females with no EOC cancer) recruited at the same regional centre, age at randomisation and year at recruitment.

We assessed the model calibration and discrimination of the predicted five-year risks. Females older than 74 years at entry were excluded. Cases that developed EOC beyond five years were treated as unaffected. For controls with a less than five-year follow-up, we predicted the EOC risks to the age at censoring. For all other controls and cases, we predicted five-year risks.

To assess model calibration, we partitioned the weighted sample into quintiles of predicted risk. Within each quintile, we compared the weighted mean of predicted risk to the weighted observed incidence using the Hosmer-Lemeshow (HL) chi-squared test.[34] To assess RR calibration, the predicted and observed RRs were calculated relative to the corresponding means of risks over all quintiles. We also compared the expected (E) with the observed (O) EOC risk within the prediction interval by calculating the ratio of expected to observed cases (E/O). The 95% confidence interval (CI) for the ratio was calculated assuming a Poisson distribution.[35]

We assessed the model discrimination between females who developed and did not develop EOC within five years using the area under the ROC curve (AUC) (Supplementary Methods).

## RESULTS

### Model description

*RAD51D, RAD51C* and *BRIP1*, based on the assumed allele frequencies and RRs, account for 2.5% of the overall model polygenic variance. Figure 1 shows the predicted EOC risks for carriers of PVs in *BRCA1, BRCA2, RAD51D, RAD51C* and *BRIP1* for various FH scenarios. With unknown FH, the risks for carriers of PVs *in RAD51D, RAD51C* and *BRIP1* are 13%, 11% and 6%, respectively. For example, for a *BRIP1* PV carrier, the risk varies from 6% for a female without EOC FH to 18% for a female with two affected first-degree relatives. The model can also be used to predict risks in families in which PVs are identified but where other family members test negative (Figure s1). For females with a FH of EOC, the reduction in EOC risk after negative predictive testing is greatest for *BRCA1* PVs, with the risks being close to (though still somewhat greater than) population risk. This effect was most noticeable for females with a strong FH. Although a risk reduction is also seen for females whose mother carried a PV in *BRCA2, RAD51D, RAD51C* or *BRIP1*, the reduction is less marked. As expected, the predicted risks are still elevated compared to population.

**Figure 1.**
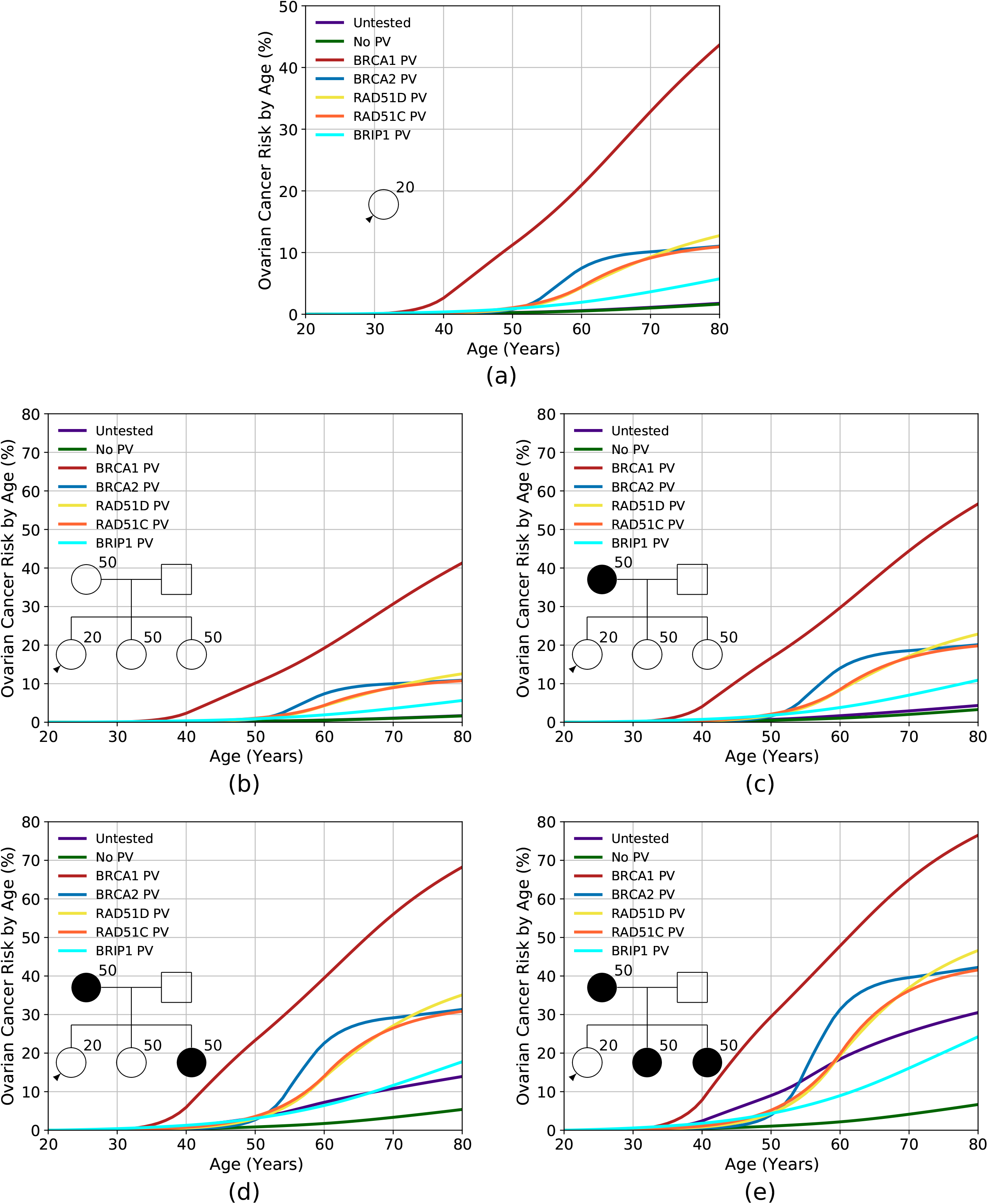
Predicted lifetime (age 20 to 80 years) EOC risk by PV and family history. Each Figure shows the risks assuming the female is untested, has no PVs, or carries a PV in *BRCA1, BRCA2, RAD51D, RAD51C* or *BRIP1*. Figure (a) assumes unknown family history. Figures (b)-(e) assume an increasing number of affected first-degree relatives, as indicated by the pedigree diagram inserts. Predictions are based on UK EOC population incidence.

Figures 2 and s2 show distributions of lifetime risk and risk by age 50 respectively for females untested for PVs, based on RFs and PRS, for two FH scenarios: (1) unknown FH (i.e. equivalent to a female from the general population); and (2) having a mother diagnosed with EOC at age 50. Table 2 shows the corresponding proportion of females falling into different risk categories. The variation in risk is greatest when including both the RFs and PRS. When considered separately, the distribution is widest for the RFs. Using the RFs and PRS combined, predicted lifetime risks vary from 0.5% for the 1^st^ percentile to 4.6% for the 99^th^ for a female with unknown FH and from 1.9% to 10.3% for a female with an affected mother.

**Table 2.** Percentage of females falling in different risk categories by status of PV in one of the high- or intermediate-risk genes included in the model and family history of cancer. The “population” risk category is defined as lifetime risk of <5% and risk to age 50 of <3%. “Moderate” risk category is defined as a lifetime risk of 5% or greater but less than 10% and a risk to age 50 of 3% or greater but less than 5%. “High” risk category is defined as a lifetime risk of 10% or greater and a risk to age 50 of 5% or greater. RF: epidemiological risk factors; PRS: polygenic risk score; NA: unknown family history; M 50: mother diagnosed with EOC at age 50. The population lifetime risk is 1.8%, and the population risk to age 50 is 0.27%.

| PV status | Family history | Risk Categories | Lifetime Risk |  |  | Risk to age 50 |  |  | Ref Fig. |
| --- | --- | --- | --- | --- | --- | --- | --- | --- | --- |
|  |  |  | RF | PRS | RF and PRS | RF | PRS | RF and PRS |  |
| Untested | NA | population | 99.9 | 100.0 | 99.4 | 100.0 | 100.0 | 100.0 | 2(a)(c) and s2(a)(c) |
|  |  | moderate | 0.1 | 0.0 | 0.6 | 0.0 | 0.0 | 0.0 |  |
|  |  | high | 0.0 | 0.0 | 0.0 | 0.0 | 0.0 | 0.0 |  |
|  | M 50 | population | 59.1 | 61.8 | 61.7 | 100.0 | 100.0 | 100.0 | 2(b)(d) and s2(b)(d) |
|  |  | moderate | 40.3 | 38.2 | 37.1 | 0.0 | 0.0 | 0.0 |  |
|  |  | high | 0.6 | 0.0 | 1.2 | 0.0 | 0.0 | 0.0 |  |
| BRCA1 | NA | population | 0.0 | 0.0 | 0.0 | 0.0 | 0.0 | 0.3 | 3(a) and s3(a) |
|  |  | moderate | 0.0 | 0.0 | 0.0 | 1.5 | 0.1 | 4.1 |  |
|  |  | high | 100.0 | 100.0 | 100.0 | 98.5 | 99.9 | 95.6 |  |
|  | M 50 | population | 0.0 | 0.0 | 0.0 | 0.0 | 0.0 | 0.0 | 3(b) and s3(b) |
|  |  | moderate | 0.0 | 0.0 | 0.0 | 0.0 | 0.0 | 0.2 |  |
|  |  | high | 100.0 | 100.0 | 100.0 | 100.0 | 100.0 | 99.8 |  |
| BRCA2 | NA | population | 1.6 | 0.1 | 4.7 | 100.0 | 100.0 | 99.9 | 3(c) and s3(c) |
|  |  | moderate | 42.8 | 37.7 | 42.3 | 0.0 | 0.0 | 0.1 |  |
|  |  | high | 55.6 | 62.2 | 52.9 | 0.0 | 0.0 | 0.0 |  |
|  | M 50 | population | 0.0 | 0.0 | 0.1 | 95.9 | 99.9 | 95.3 | 3(d) and s3(d) |
|  |  | moderate | 1.9 | 0.1 | 4.7 | 4.0 | 0.1 | 4.5 |  |
|  |  | high | 98.1 | 99.9 | 95.3 | 0.0 | 0.0 | 0.2 |  |

| PV | Family | Risk | Lifetime Risk |  |  | Risk to age 50 |  |  | Ref Fig. |
| --- | --- | --- | --- | --- | --- | --- | --- | --- | --- |
| **RAD51D** | NA | population | 0.6 | 0.0 | 2.1 | 100.0 | 100.0 | 99.8 | 3(e)<br>and<br>s3(e) |
|  |  | moderate | 24.5 | 18.3 | 30.8 | 0.0 | 0.0 | 0.2 |  |
|  |  | high | 75.0 | 81.7 | 67.1 | 0.0 | 0.0 | 0.0 |  |
|  | M 50 | population | 0.0 | 0.0 | 0.0 | 93.5 | 99.6 | 92.6 | 3(f)<br>and<br>s3(f) |
|  |  | moderate | 0.6 | 0.0 | 2.0 | 6.3 | 0.4 | 7.0 |  |
|  |  | high | 99.4 | 100.0 | 98.0 | 0.2 | 0.0 | 0.4 |  |
| **RAD51C** | NA | population | 1.8 | 0.1 | 4.9 | 99.8 | 100.0 | 99.3 | 3(g)<br>and<br>s3(g) |
|  |  | moderate | 44.4 | 39.3 | 43.1 | 0.2 | 0.0 | 0.7 |  |
|  |  | high | 53.7 | 60.6 | 52.0 | 0.0 | 0.0 | 0.0 |  |
|  | M 50 | population | 0.0 | 0.0 | 0.1 | 85.8 | 96.2 | 85.0 | 3(h)<br>and<br>s3(h) |
|  |  | moderate | 2.0 | 0.1 | 5.0 | 13.6 | 3.8 | 13.7 |  |
|  |  | high | 98.0 | 99.9 | 95.0 | 0.6 | 0.0 | 1.3 |  |
| **BRIP1** | NA | population | 43.6 | 34.0 | 45.7 | 100.0 | 100.0 | 99.7 | 3(i) and<br>s3(i) |
|  |  | moderate | 52.3 | 65.0 | 47.4 | 0.0 | 0.0 | 0.3 |  |
|  |  | high | 4.1 | 1.0 | 6.9 | 0.0 | 0.0 | 0.0 |  |
|  | M 50 | population | 2.1 | 0.1 | 4.8 | 93.0 | 99.3 | 91.4 | 3(j) and<br>s3(j) |
|  |  | moderate | 43.5 | 38.9 | 44.5 | 6.8 | 0.7 | 8.1 |  |
|  |  | high | 54.5 | 61.1 | 50.7 | 0.3 | 0.0 | 0.5 |  |

**Figure 2.**
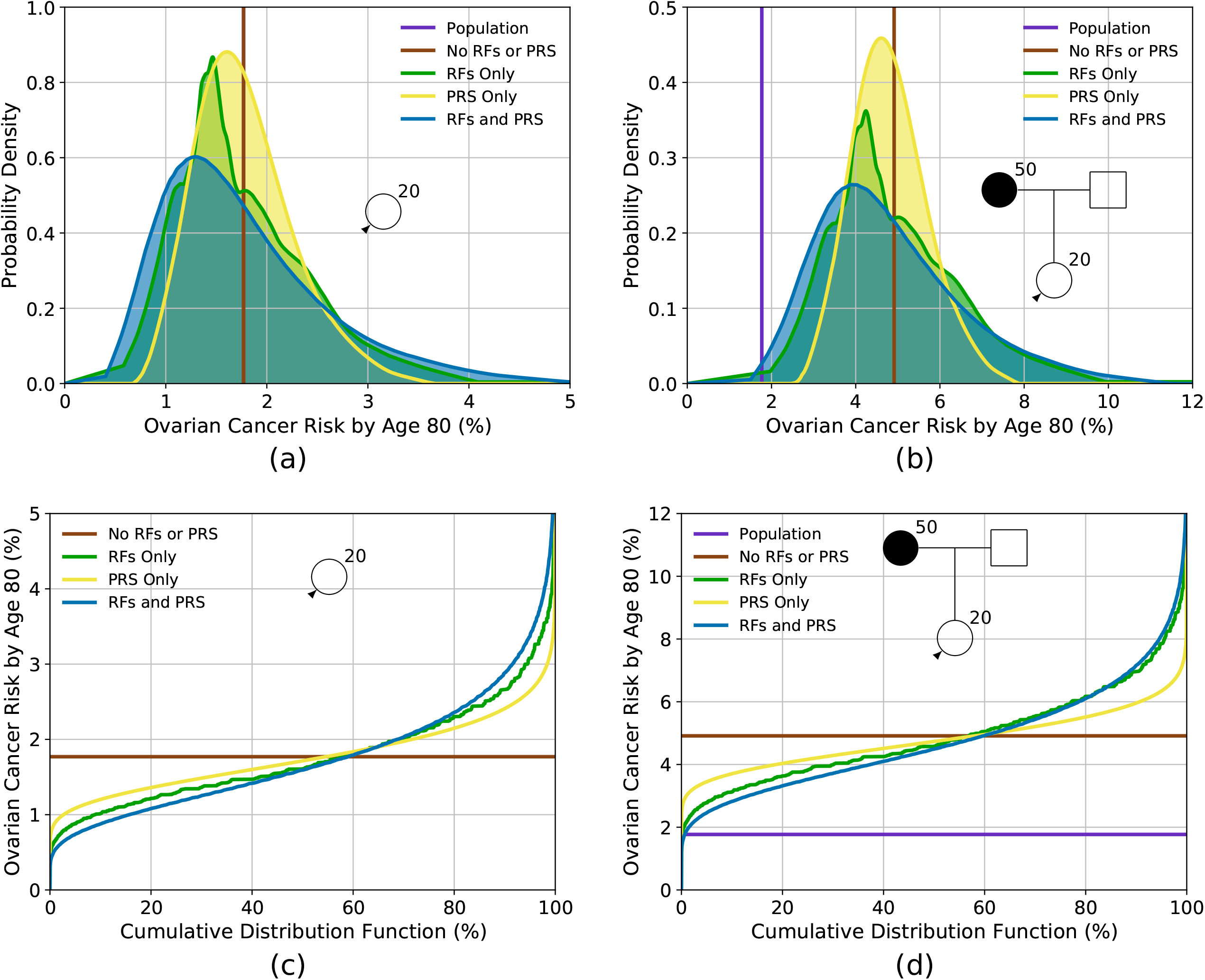
Predicted lifetime (age 20 to 80 years) EOC risk for a female untested for PVs based on the different predictors of risk (risk factors (RFs) and PRS). Figures (a) and (c) show the risk for a female with an unknown family history (equivalent to the distribution of risk in the population), while figures (b) and (d) show the risk for a female with a mother affected at age 50. Figures (a) and (b) show the probability density function against absolute risk, while figures (c) and (d) show absolute risk against cumulative distribution. The vertical line in figure (a) and the horizontal line in Figure (c) (labelled “no RFs or PRS”) is equivalent to the population risk of EOC. The “Population” risk is shown separately in figures (b) and (d). Predictions are based on UK EOC population incidences.

Figure 3 shows the predicted lifetime EOC risk for carriers of PVs in *BRCA1, BRCA2, RAD51D, RAD51C* and *BRIP1* based on RFs and PRS for two FH scenarios. Taking a *RAD51D* PV carrier, for example, based on PV testing and FH alone, the predicted risks are 13% when FH is unknown and 23% when having a mother diagnosed with EOC at age 50. When RFs and the PRS are considered jointly, risks vary from 4% for those at the 1^st^ percentile to 28% for the 99^th^ with unknown FH and from 9% to 43% with an affected mother. Table 1 shows the proportion of females with PVs falling into different risk categories. Based on the combined distribution, 33% of *RAD51D* PV carriers in the population are expected to have a lifetime EOC risk of less than 10%. Similarly, the distributions of risk for *BRIP1* PV carriers are shown in Figure 3(i) and 3(j) and in Table 1. Based on the combined RFs and PRS distributions, 46% of *BRIP1* PV carriers in the population are expected to have lifetime risks of less than 5%; 47% to have risks between 5% and 10%, and 7% to have risks of 10% or greater. A BRIP1 PV carrier with an affected mother, on the basis of FH alone, has a lifetime risk of 11%. However, when the RFs and PRS are considered, 50% of those would be reclassified as having lifetime risks of less than 10%.

**Figure 3.**
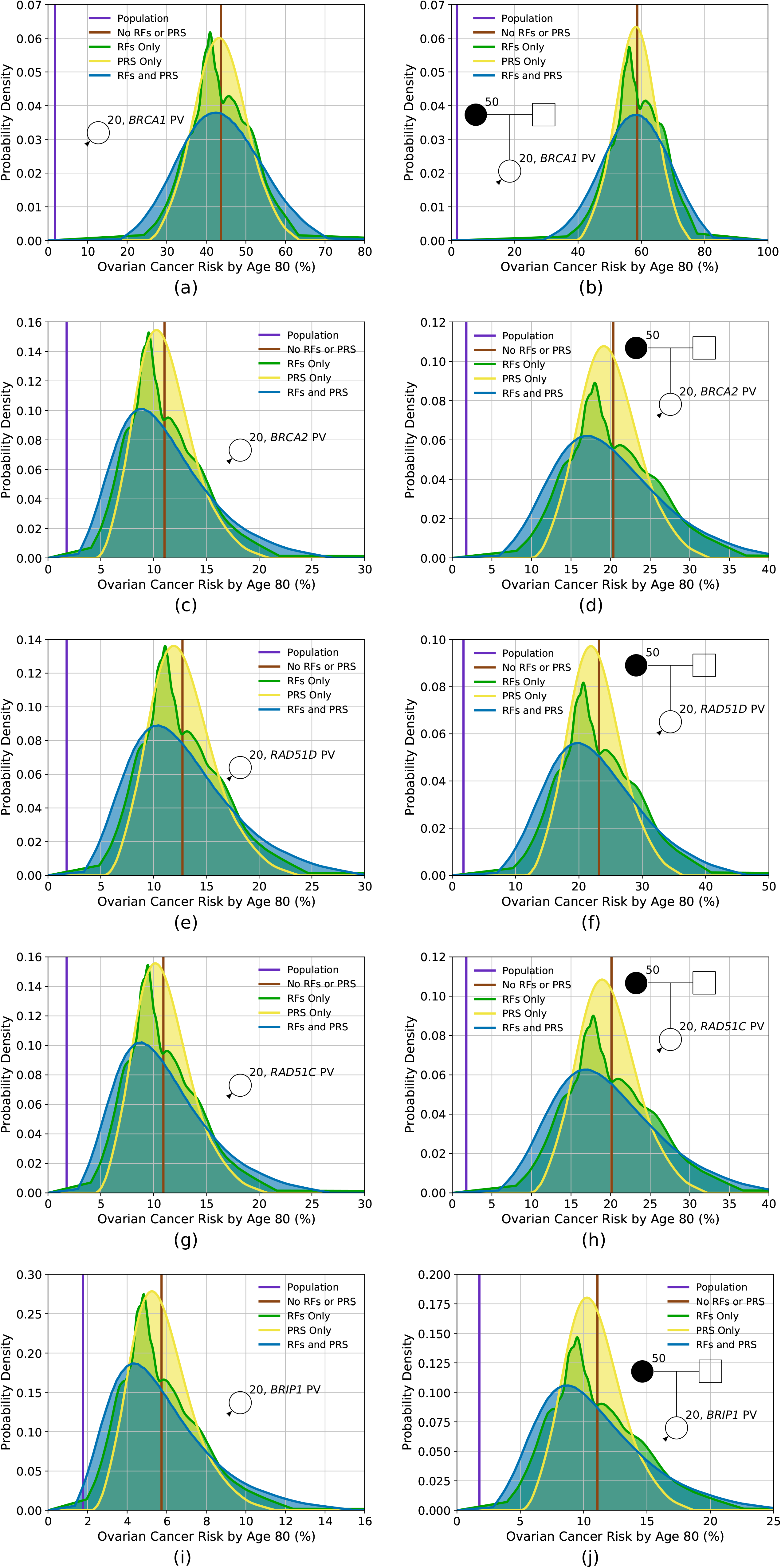
Predicted lifetime EOC risk for a female who has a PV in one of the high- or intermediate-risk genes included in the model, based on the different predictors of risk (risk factors (RFs) and PRS), for two family histories. Figures (a) and (b) show the lifetime risk for a carrier of a PV in *BRCA1*, Figures (c) and (d) show the lifetime risk for a carrier of a PV in *BRCA2*, Figures (e) and (f) show the lifetime risk for a carrier of a PV in *RAD51D*, Figures (g) and (h) show the lifetime risk for a carrier of a PV in *RAD51C*, while Figure (i) and (j) show the lifetime risk for a carrier of a PV in *BRIP1*. Figures (a), (c), (e), (g) and (i) show risks for an unknown family history, while Figures (b), (d), (f), (h) and (j) show risks for a female whose mother is diagnosed with EOC at age 50. Predictions based on UK ovarian cancer incidences.

Figures s4 and s5 show the probability trees describing the reclassification of females as more information (RFs, PRS, and testing for PVs in the MGs) is added to the model for a female with unknown FH and a female with a mother diagnosed at age 50 respectively based on the predicted lifetime risks. Figures s4(a) and s5(a) show the reclassification resulting from adding RFs, MG and PRS sequentially, while Figures s4(b) and s5(b) assume the order RFs, PRS and then MG. Assuming the three risk categories for lifetime risks are <5%, ≥5% but <10% and ≥10%, there is significant reclassification as more information is added.

### Model validation

After censoring, 1961 participants with 374 incident cases and 1587 controls met the 5-year risk prediction eligibility criteria. Table s5 summarises their characteristics at baseline.

The model considering FH, the 15-variant PRS and a subset of the RFs (but not including testing for PVs in the MGs) demonstrated good calibration in both absolute and relative predicted risk (Figure 4). Over the five-year period, the model predicted 391 EOCs, close to the 374 observed (E/O=1.05, 95%CI=0.94-1.16). The model was well calibrated across the quintiles of predicted risk (HL p=0.08), although there was a suggestion for an underprediction of risk in the lowest quintile (absolute risk E/O=0.66, 95%CI=0.52-0.91; RR E/O=0.63, 95%CI:0.42-0.95). The AUC for assessing discrimination of these model components was 0.61 (95%CI:0.58-0.64).

**Figure 4.**
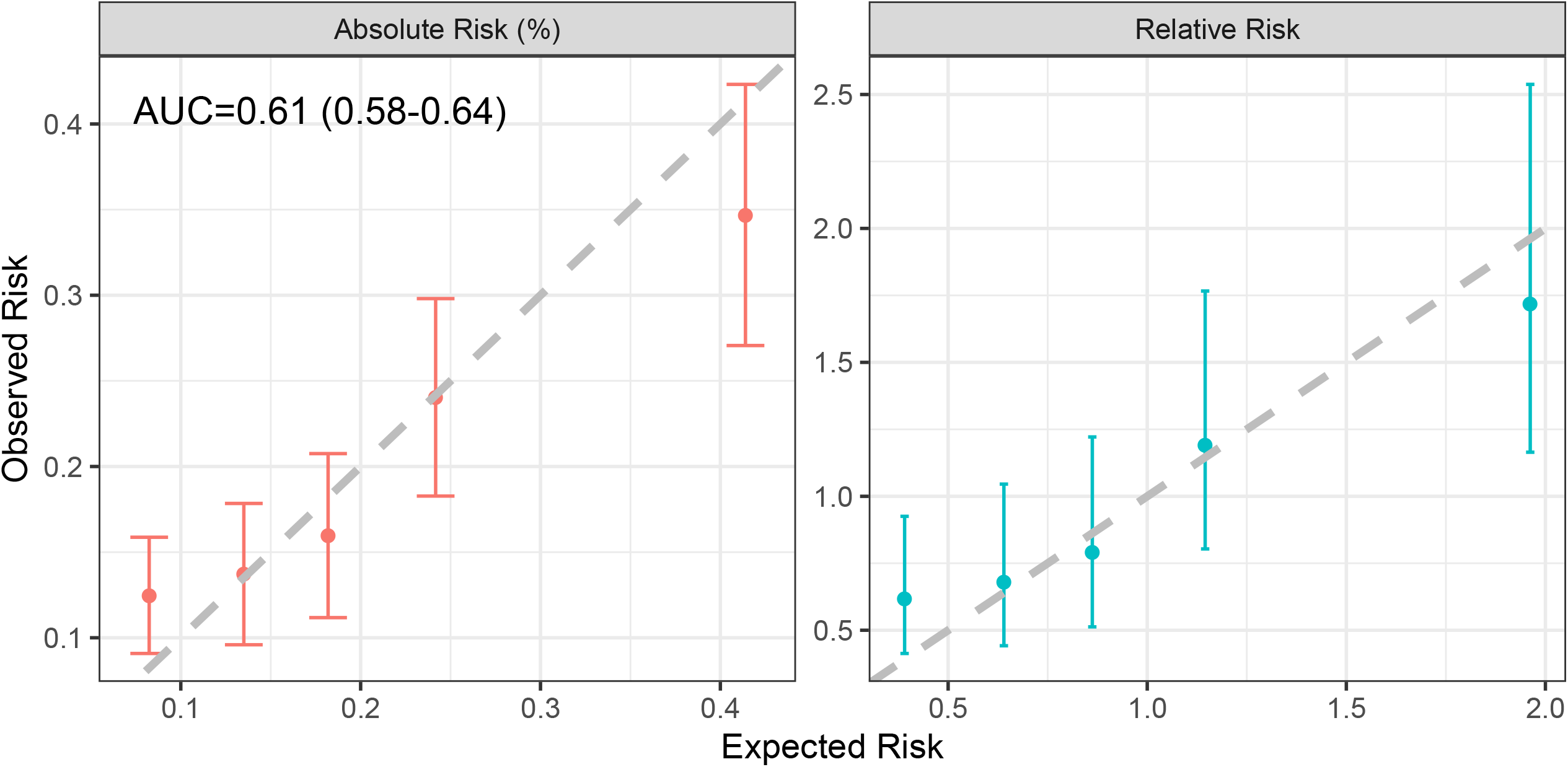
Calibration of the absolute and relative predicted five-year EOC risks, showing the observed and expected risks by quintile. The bars show the 95% confidence intervals for the observed risks. Relative risks were calculated relative to the overall mean of observed and predicted risks.

When looking at individual factors, FH predicted the widest 5-year risk variability (sd=0.0013; range 0.04% to 4.0%), followed by RFs (sd=0.0010; range:0.02%-0.7%) and PRS (sd=0.0009; range:0.05%-1.0%, Figure s6). As expected, their sequential inclusion increased the variability (sd=0.0018; Figure s6).

## DISCUSSION

The EOC risk prediction model presented here combines the effects of FH, the explicit effects of rare moderate- to high-risk PVs in five established EOC susceptibility genes, a 36-variant PRS and other clinical and epidemiological factors (Table 1). The model provides a consistent approach for estimating EOC risk on the basis of all known factors and allows for prevention approaches to be targeted at those at highest risk.

The results demonstrate that in the general population (unknown FH), the existing PRS and RF alone identify 0.6% of females who have a lifetime risk of >5% (Table 2). On the other hand, for females with FH, 37.1% of females would have a predicted risk between 5% and 10% and 1.2% would have an EOC risk of ≥10% (Table 2). The results show that the RFs provide a somewhat greater level of risk stratification than the 36-variant PRS. However, discrimination is greater when both are considered jointly. These results were in line with the observed risk distributions in the validation dataset, but direct comparisons were not possible due to the different variants included in the PRSs and limited RFs in the validation study. The results also show that significant levels of risk re-categorisation can occur for carriers of PVs in moderate or high-risk susceptibility genes.

The comprehensive risk model is based on a synthetic approach previously used for BC[28] and makes several assumptions. In particular, we assumed that the risks associated with known RFs and the PRS combine multiplicatively. We have not assessed this assumption in the present study, however, published studies found no evidence of deviations from the multiplicative model for the combined effect of the RFs and the PRS,[26] suggesting that this assumption is reasonable. The model assumes that the RFs are also independent of the residual polygenic component that captures the effect of FH. However, for the RFs included, we used estimates from published studies that have adjusted for the other known EOC RFs. The observation that the model was calibrated on the RR scale in the UKCTOCS validation study also suggests that these assumptions are broadly valid.

Similarly, the model assumes that the relative effect-sizes of RFs and the PRS are similar in females carrying PVs in *BRCA1, BRCA2, RAD51C, RAD51D* and *BRIP1* and those without PVs in these genes. Evidence from studies of *BRCA1* and *BRCA2* PV carriers suggests that this assumption is plausible: PRSs for EOC have been shown to be associated with similar RRs in the general population and in *BRCA1* and *BRCA2* PV carriers.[32, 36, 37] The current evidence also suggests that known RFs have similar effect sizes in *BRCA1* and *BRCA2* PV carriers as in non-carriers.[38, 39] No studies have so far assessed the joint effects of *RAD51C, RAD51D* and *BRIP1* PVs with the PRS, but the observation that FH modifies EOC risk for *RAD51C/D* PV carriers[27] suggests that similar arguments are likely to apply. Large prospective studies are required to address these questions in more detail. We were not able to validate these assumptions explicitly in UKCTOCS because gene-panel testing data were not available.

Other RFs for EOC that have been reported in the literature include breast feeding[40] and age at menarche and menopause.[23] However, the evidence for these RFs is still limited. Our model is flexible enough to allow for additional RFs to be incorporated in the future.

We validated the five-year predicted risks on the basis of FH, RFs and PRS available in an independent dataset from a prospective trial.[2] A key strength was that EOC was a primary outcome in UKCTOCS. All cases were reviewed and confirmed by an independent outcome review committee.[2] The results indicated that absolute and RRs were well-calibrated overall and in the top quintiles of predicted risk. However, there was some underprediction of EOC in the bottom quintile. This could be due to differences in the RF distributions in those who volunteer to participate in research (self-selected more healthy individuals[41]) compared to the general population or due to random variations in the effects of the RFs in UKCTOCS compared to other studies. Alternatively, the multiplicative assumption may break down in the lowest risk category. Further large prospective cohorts will be required to determine whether the underprediction in the lowest risk category reflects a systematic miscalibration of the model or is due to chance. Although the AUC based on model components in this validation study was modest, it is not surprising given that only a subset of the model predictors were used, and UKCTOCS recruited primarily low-risk women. Inclusion of the optimal PRS,[32] all RFs and information on PVs in the five genes that account for a large fraction of the EOC FRR are expected to lead to an increase in AUC.

The current validation study has some limitations. The underlying model accounts for FH information on both affected and unaffected family members, but the UKCTOCS recruitment questionnaire did not include information on unaffected family members. Family sizes and ages for unobserved family members were imputed using demographic data. In addition, since data on whether the affected family members were from the paternal or maternal side, we assumed all the affected family members were from the same (maternal) side. These may result in inaccuracies in risk predictions. A further limitation is that UKCTOCS was undertaken to assess screening of low-risk women and therefore is not necessarily representative of a true population cohort, as females with a FH of two or more relatives with EOC or who were known carriers of *BRCA1/2* PVs were not eligible to participate in the randomised controlled trial. Data were not available on the rare moderate and high-risk PVs, and we were only able to assess a PRS with 15-variants, rather than the more informative 36-variant PRS. Therefore, it has not been possible to validate the full model presented here. Future analyses in other cohorts will be required to further validate the full model.

In summary, we have presented a methodological framework for a comprehensive EOC risk prediction model that considers the currently known genetic and epidemiologic RFs and explicit FH. The model allows users to obtain consistent, individualised EOC risks. It can also be used to identify target populations for studies to assess novel prevention strategies (such as salpingectomy) or early detection approaches by identifying those at higher risk of developing the disease for enrolment into such studies. Future independent studies should aim to validate the full model, including the full PRS and rare PVs in diverse settings. The model is available via the CanRisk Tool (www.canrisk.org), a user-friendly web tool that allows users to obtain future risks of developing EOC.

## Data Availability

The model is freely available at www.canrisk.org.
For access to UKCTOCS data set, which is subject to GDPR rules, please contact the UKCTOCS Biobank coordinator. The data access process is outlined here http://uklwc.mrcctu.ucl.ac.uk/access-process/

## Acknowledgements

This work has been supported by grants from Cancer Research UK (C12292/A20861).

The analysis is part of PROMISE, which was funded through Cancer Research UK PRC Programme Grant A12677 and by The Eve Appeal. University College London investigators received support from the National Institute for Health Research University College London Hospitals Biomedical Research Centre and from MRC core funding (MR_UU_12023). UKCTOCS was core funded by the Medical Research Council (G9901012 and G0801228), Cancer Research UK (C1479/A2884), and the Department of Health with additional support from the Eve Appeal.

FMW was supported by a National Institute for Health Research (NIHR) Clinician Scientist award (NIHR-CS-012-03).

MT was funded by the European Union Seventh Framework Program (2007– 2013)/European Research Council (310018).

The authors are particularly grateful to those throughout the UK who are participating in the trial and to the centre leads and the entire medical, nursing and administrative staff who work on the UKCTOCS.

## Ethics Declaration

Ethics approval UKCTOCS was approved by the UK North West Multicentre Research Ethics Committees (North West MREC 00/8/34) on 21 June 2000 with site-specific approval from the local regional ethics committees and the Caldicott guardians (data controllers) of the primary care trusts. The SNP protocol was approved by NRES Committee North West - Liverpool Central (14/NW1026) in June 2014.

## Data availability

The model is freely available at www.canrisk.org. For access to UKCTOCS data set, which is subject to GDPR rules, please contact the UKCTOCS Biobank coordinator. The data access process is outlined here http://uklwc.mrcctu.ucl.ac.uk/access-process/

## Author Contributions

Conceptualization: ACA, PPDP, DFE

Methodology: AL, XY, ACA, DFE, PPDP, JT

Software: AL, XY, APC, TC, SA

Investigation: AL, XY, ACA, PPDP, DFE, SAG, SJR, FG, RM, FWM, MT

Data curation: AGM, AR, GL

Formal analysis: AL, XY, JT, NM

Funding acquisition: ACA, IJ, UM

Resources: PPDP, AGM, AR, JT, SAG, SJR, IJ, UM

Supervision: ACA

Visualisation: AL, XY

Writing – original draft: AL, XY, ACA

Writing – review & editing: all authors

