## Supplementary Material for "A Comprehensive Epithelial Tubo-Ovarian Cancer Risk Prediction Model Incorporating Genetic and Epidemiological Risk Factors"

#### Model Extensions

The model has been extended to incorporate the effects of high- and intermediate-risk pathogenic variants (PV) in RAD51D, RAD51D and BRIP1, the effects of a polygenic risk score (PRS) and epidemiological risk factors (RF).

##### Polygenic Risk Scores

The model has been extended to incorporate the effects of common variants, summarised in a polygenic risk score (PRS). Here we detail how to calculate a polygenic risk score from an individual's genotypes and the set of variants used in the PRS.

To calculate a person's PRS, we use the framework described in [1]. For a set of $N$ common variants, labelled $i = 1, \dots, N$, each with log relative risk $\beta_i$, we calculate an individual's raw PRS as

$$PRS = \sum_{i=1}^{N} D_i \beta_i,$$

where $D_i$ is the person's genetic dosage for variant $i$, which is in the range $[0, 2]$. The dosage may come directly from the person's genotype or may be imputed.

The variance explained by this PRS is

$$\sigma_{PRS}^2 = \sum_{i=1}^{N} \sigma_i^2,$$

where $\sigma_i^2$ is the variance for variant $i$, given by

$$\sigma_i^2 = log\left(\frac{(1-f_i)^2 + 2(1-f_i)f_i e^{2\beta_i} + f_i^2 e^{4\beta_i}}{\left((1-f_i)^2 + 2(1-f_i)f_i e^{\beta_i} + f_i^2 e^{2\beta_i}\right)^2}\right),$$

where $f_i$ is the effect allele frequency.

The mean PRS is given by

$$\mu_{PRS} = \sum_{i=1}^{N} 2 f_i \beta_i.$$

The model takes as input the standard normal PRS (z-score), given by

$$PRS_z = (PRS - \mu_{PRS})/\sigma_{PRS},$$

and the square root of the overall polygenic variance in the model explained by the PRS is

$$\alpha = \frac{\sigma_{PRS}}{1.4156}.$$

In [2] the polygenic variance was determined to be 1.434, which implicitly included the effects of *RAD51D*, *RAD51C* and *BRIP1*. *RAD51D*, *RAD51C* and *BRIP1* are explicitly included in this version of the model, and so their effects must be subtracted from the polygenic variance, leaving 1.4156.

The above procedure allows the set of variants to change between people, for instance, people may only have been genotyped for a subset of the known variants (e.g., an older PRS or missing genotypes), or more variants may be added.

| Variant Name | Chromosome | Position | Reference Allele | Effect Allele | Effect Allele Frequency | Log Odds Ratio |
|---|---|---|---|---|---|---|
| rs3820282 | 1 | 22468215 | C | T | 0.155486 | 0.0808 |
| rs12039431 | 1 | 38082122 | G | A | 0.254292 | 0.0835 |
| rs2165109 | 2 | 111818658 | A | C | 0.252134 | 0.0642 |
| rs1470053 | 2 | 111915946 | G | T | 0.186134 | -0.0118 |
| rs895412 | 2 | 113973964 | T | C | 0.464965 | 0.0518424 |
| rs72831810 | 2 | 113979364 | G | A | 0.149592 | 0.0322626 |
| rs1318778 | 2 | 177037831 | C | G | 0.680296 | -0.1005 |
| rs62276623 | 3 | 156402487 | C | T | 0.0494519 | 0.3647 |
| rs9869209 | 3 | 190531882 | G | A | 0.303218 | -0.0668 |
| rs34902361 | 4 | 70577859 | G | A | 0.351856 | -0.0574 |
| rs10069690 | 5 | 1279790 | C | T | 0.258965 | 0.035113479 |
| rs7705526 | 5 | 1285974 | C | A | 0.330049 | 0.058960178 |
| rs2853677 | 5 | 1287194 | G | A | 0.570409 | -0.084927779 |
| rs2853669 | 5 | 1295349 | A | G | 0.306276 | -0.079717471 |
| rs336126 | 5 | 54476556 | G | A | 0.730798 | -0.0666 |
| rs11782652 | 8 | 82653644 | A | G | 0.0678244 | 0.1294 |
| rs9886651 | 8 | 128817883 | A | G | 0.456948 | 0.0759846 |
| rs35916594 | 8 | 129069820 | G | A | 0.375132 | -0.0692237 |
| rs6470611 | 8 | 129217984 | G | C | 0.477535 | 0.048478 |
| rs10088755 | 8 | 129551633 | G | A | 0.130493 | -0.175939 |
| rs62543619 | 9 | 16914716 | G | A | 0.204783 | -0.139195 |
| rs10810671 | 9 | 16914835 | A | C | 0.322426 | -0.101013 |
| rs9406757 | 9 | 19044489 | G | A | 0.136561 | 0.0788 |
| rs10739885 | 9 | 106912892 | G | A | 0.547349 | 0.0581903 |
| rs635634 | 9 | 136155000 | C | T | 0.196517 | 0.0931 |
| rs7084454 | 10 | 21821274 | G | A | 0.331184 | 0.0799 |
| rs71479294 | 10 | 112011084 | A | G | 0.148067 | 0.0752 |
| rs7139079 | 12 | 121415293 | G | A | 0.579263 | -0.0593 |
| rs76119208 | 15 | 91535329 | G | T | 0.133133 | -0.08040702 |
| rs11657964 | 17 | 36100767 | A | G | 0.602031 | -0.0582 |
| rs169201 | 17 | 44790203 | A | G | 0.201454 | 0.102556 |
| rs12946636 | 17 | 46472432 | C | G | 0.272844 | 0.118166 |
| rs10853591 | 18 | 21425852 | T | C | 0.61887 | -0.0283 |
| rs4808075 | 19 | 17390291 | T | C | 0.296534 | 0.0796744 |
| rs12982058 | 19 | 17409380 | C | T | 0.51289 | -0.06128376 |
| rs2070368 | 21 | 36080398 | T | C | 0.406715 | -0.0599 |

*Table s1. List of 36 common variants developed by the Ovarian Cancer Association Consortium [3].*

#### Parameterisation of the Risk Factors

The model has been extended to incorporate the effects of epidemiological risk factors. In the model, risk factors are parameterised by their population distribution and relative risk, given in Table s2.

| ***Risk Factor*** | ***Population Distribution*** | ***Relative Risk*** | | **Reference for Distribution** | **Reference for Relative Risk** |
|---|---|---|---|---|---|
| **Parity** | | **<20** | **≥20** | | |
| *0* | 0.28 | 1.0 | 1.00 | [4] from [5] Table 3 | [6] Table 4 |
| *1* | 0.16 | 1.0 | 0.76 | | |
| *>1* | 0.56 | 1.0 | 0.58 | | |
| **Oral Contraceptive Use (years of use)** | | **<current age** | **≥current age** | | |
| *Never or <1* | 0.710 | 1.0 | 1.00 | [7] Table 2 | [7] Table 2 |
| *1-4* | 0.138 | 1.0 | 0.78 | | |
| *5-9* | 0.086 | 1.0 | 0.64 | | |
| *10-14* | 0.046 | 1.0 | 0.56 | | |
| *≥15* | 0.020 | 1.0 | 0.42 | | |
| **MHT Use** | | **<current age** | **≥current age** | | |
| *Never* | 0.73 | 1.0 | 1.00 | [8] Table 2 | [9] Abstract |
| *Ever* | 0.27 | 1.0 | 1.37 | | |
| **Tubal Ligation** | | **<20** | **≥20** | | |
| *No* | 0.77 | 1.0 | 1.00 | [10] Table 1 | [6] Table 4 |
| *Yes* | 0.23 | 1.0 | 0.74 | | |
| **Endometriosis** | | **<20** | **≥20** | | |
| *No* | 0.90 | 1.0 | 1.00 | [11] Abstract | [6] Table 4 |
| *Yes* | 0.10 | 1.0 | 1.53 | | |
| **BMI** | | **<20** | **≥20** | | |
| *<22.5* | 0.325 | 1.0 | 1.00 | [12] Table 2 | [12] Table 2 |
| *[22.5, 30)* | 0.529 | 1.0 | 1.066 | | |
| *≥30* | 0.146 | 1.0 | 1.13 | | |
| **Height (cm)** | | **<20** | **≥20** | | |
| *<152.91* | 0.0625 | 1.0 | 0.82 | [4] from [13, 14] | [15] Abstract |
| *[152.91, 159.65)* | 0.25 | 1.0 | 0.91 | | |
| *[159.65, 165.96)* | 0.375 | 1.0 | 1.00 | | |
| *[165.96, 172.70)* | 0.25 | 1.0 | 1.10 | | |
| *≥172.70* | 0.0625 | 1.0 | 1.22 | | |

*Table s2. Summary of the EOC parameterisations of the risk factors used in the model. For oral contraceptive use and menopause hormone therapy (MHT) use the relative risks are taken to be 1.0 up to the proband's current age.*

#### Parameterisation of the Rare Variants

The effects of pathogenic variants are parameterised in the model via their allele frequency and relative risk, give in Table s3.

| GENE | ALLELE FREQUENCY | RELATIVE RISK | | SOURCE |
|---|---|---|---|---|
| ***RAD51D*** | 0.00026 | $1$<br>$\exp(-2.88662 + 0.09656 \times \text{age})$<br>$\exp(\ \ 5.99144 - 0.05651 \times \text{age})$ | $age < 30$<br>$30 \leq age < 58$<br>$age \geq 58$ | [16] |
| ***RAD51C*** | 0.00022 | $1$<br>$\exp(-1.7974 + 0.07631 \times \text{age})$<br>$\exp(\ \ 9.7592 - 0.1163 \times \text{age})$ | $age < 30$<br>$30 \leq age < 60$<br>$age \geq 60$ | [16] |
| ***BRIP1*** | 0.00044 | $3.41\ (2.12 - 5.54)$ | | [17] |

*Table s3. The parameters used to include the effects of rare intermediate-risk variants in the model. Risks are relative to the general population.*

#### Other Model Components

Previously [2], the model included FH of EOC and first female breast cancer (BC). To align with the BOADICEA model [18], the model was extended to take account of female contralateral BC, male BC, prostate cancer and pancreatic cancer, assuming that the relative risk for carriers of pathogenic variants (PVs) in *BRAC1* and *BRCA2* is the same as that in the BOADICEA model [19]. We assumed that PVs in *RAD51D*, *RAD51C* and *BRIP1* do not increase the risks for these cancers relative to the population.

Further, using the methodology developed in [20] we included the effects of tumour pathology subtype of a first BC for females, where we assumed that the pathology proportions for carriers of PVs in *BRAC1* and *BRCA2* are the same as those in the BOADICEA model [21] and that the pathology proportions for carriers of PVs in *RAD51D*, *RAD51C* and *BRIP1* are the same as those in the general population. FH of these cancers and first BC pathology can be indicative of PVs in *BRAC1* or *BRCA2*.

### Model Validation

#### Study Subjects

The United Kingdom Collaborative Trial of Ovarian Cancer Screening (UKCTOCS) is a randomised controlled trial for assessing the effect of screening on EOC mortality initiated in 2001 [22, 23]. Postmenopausal females aged 50-74 years were invited to participate. Participants provided a blood sample and completed a baseline questionnaire that included information on personal and family cancer history, number of pregnancies lasting at least 6 months, OCP, MHT, sterilisation, height and weight. Information on endometriosis was not collected. All participants provided written informed consent. Females were excluded if they were at increased risk of EOC due to family history of breast or ovarian cancer or if they were known carriers of EOC predisposing PVs, or had self-reported previous bilateral oophorectomy or ovarian malignancy or active non-ovarian malignancy. Two follow-up questionnaires were administered, the first 3-5 years post-randomisation and the second in 2014 [23, 24]. Notification of cancer diagnoses and deaths were through NHS Digital for the females residing in England and the Northern Ireland Cancer Registry and Central Services Agency for those residing in Northern Ireland. For females who developed EOC, medical notes were retrieved and independently reviewed by an Outcomes Review Committee who assigned histological subtype, stage and grade.

For the present study, a nested case-control design was adopted. Cases were defined as females diagnosed with incident invasive epithelial ovarian or fallopian tube cancers or primary peritoneal cancer. To exclude potentially prevalent cases at recruitment, we predicted the risk of EOC from the age at recruitment "plus one year" and excluded samples with follow-up time less than one year. Two random controls were selected per case, matched on regional centre, age at randomisation and year at recruitment [24]. Participants who had a previous cancer diagnosis except for breast and non-melanoma skin cancer before recruitment were excluded.

#### Model Discrimination

The model discrimination was assessed by area under the ROC curve (AUC) and Harrell's C index [25]. The AUC was estimated as the weighted probability that the predicted risks for cases outrank the risks for controls. Suppose $D$ is the EOC indicator (i.e., $D = 1$ for cases and $D = 0$ for controls), then, for any case-control pair of individuals $i$ and $j$,

$$AUC = P_{weighted}\left(P_i > P_j \middle| D_i = 1, D_j = 0\right)$$

$$= \frac{\sum_{\substack{i \in cases \\ j \in controls}} I\left(P_i > P_j\right) W_i W_j}{\sum_{\substack{i \in cases \\ j \in controls}} W_i W_j}$$

where $I$ denotes the indicator function and $W_i$ is the weight for individual $i$.

| | CONTROLS | CASES |
|---|---|---|
| **Number of females** | 1587 | 374 |
| **Age at baseline** | **N (%)** | |
| *<60* | 490 (30.9) | 105 (28.1) |
| *60-70* | 787 (49.6) | 196 (52.4) |
| *≥70* | 310 (19.5) | 73 (19.5) |
| **Mean age at baseline (sd)** | 63 (6.1) | 63 (6.0) |
| **Year of birth** | **N (%)** | |
| *<1930* | 21 (1.3) | 4 (1.1) |
| *1930-1939* | 701 (44.2) | 164 (43.9) |
| *1940-1949* | 758 (47.8) | 183 (48.9) |
| *≥1950* | 107 (6.7) | 23 (6.1) |
| **Parity** | **N (%)** | |
| *0* | 221 (13.9) | 53 (14.2) |
| *1* | 198 (12.5) | 31 (8.3) |
| *>1* | 1157 (72.9) | 288 (77) |
| *Missing* | 11 (0.7) | 2 (0.5) |
| **Oral contraceptive use (years)** | **N (%)** | |
| *Never or < 1* | 714 (45) | 189 (50.5) |
| *1-4* | 403 (25.4) | 86 (23) |
| *5-9* | 203 (12.8) | 44 (11.8) |
| *10-14* | 161 (10.1) | 38 (10.2) |
| *≥15* | 96 (6) | 17 (4.5) |
| *Missing* | 10 (0.6) | 0 (0) |
| **MHT use** | **N (%)** | |
| *Never* | 1276 (80.4) | 304 (81.3) |
| *Ever* | 311 (19.6) | 70 (18.7) |
| **Sterilisation** | **N (%)** | |
| *No* | 1285 (81) | 301 (80.5) |
| *Yes* | 302 (19) | 73 (19.5) |
| **BMI (kg/m2)** | **N (%)** | |
| *<22.5* | 295 (18.6) | 63 (16.8) |
| *22.5-30* | 976 (61.5) | 237 (63.4) |
| *≥30* | 301 (19) | 72 (19.3) |
| *Missing* | 15 (0.9) | 2 (0.5) |
| **Height (cm)** | **N (%)** | |
| *<152.91* | 165 (10.4) | 34 (9.1) |
| *[152.91, 159.65)* | 349 (22) | 96 (25.7) |
| *[159.65, 165.96)* | 693 (43.7) | 149 (39.8) |
| *[165.96, 172.70)* | 275 (17.3) | 68 (18.2) |
| *≥172.70* | 96 (6) | 27 (7.2) |
| *Missing* | 9 (0.6) | 0 (0) |
| **Non-EOC in 1st and 2nd Degree relatives, N (%)** | 1507 (95) | 344 (92) |
| **Non-BC in 1st and 2nd Degree relatives, N (%)** | 1232 (77.6) | 274 (73.3) |

*Table s4. A summary of the characteristics of the subjects at baseline and follow-up time by EOC status (N: number of individuals; sd: standard deviation; MHT: menopausal hormone therapy; BMI: body mass index).*

#### Pedigree construction

The baseline questionnaire collected information on whether the mother was diagnosed with ovarian or breast cancer and the number of daughters, number of grandmothers, number of granddaughters, number of sisters and number of aunts diagnosed with ovarian or breast cancer. Based on these, we constructed a pedigree for each case and control that included information on first- and second-degree relatives. The size of each nuclear family within each pedigree was determined by randomly sampling from the cohort-specific distribution of family sizes for the UK [26]. Using the information on the year of birth and age of the proband, reported at baseline, we randomly assigned each family member a year of birth and age at last observation under the following assumptions: (a) the age gap between successive generations ranged from 18 to 45 years with a mean age gap equal to 25 years; (b) spouses were assumed to have the same age; (c) vital status and age at last follow-up were assigned based on cohort-specific life expectancy tables for England and Wales [27]. For simplicity, we assumed that the reported breast and ovarian cancers in relatives occurred in different family members. We assumed that all affected individuals came from the maternal side and assigned an age at ovarian/breast cancer diagnosis by sampling from the cohort-specific probability based on the population incidences of ovarian or breast cancer for the UK [28].

#### Validation PRS

As the validation set had not been genotyped for all the common variants in Table s1, a PRS was constructed using the set of variants given in Table s5.

| Variant Name | Chromosome | Position | Reference Allele | Effect Allele | Effect Allele Frequency | Log Odds Ratio |
|---|---|---|---|---|---|---|
| rs58722170 | 1 | 38096421 | G | C | 0.2208 | 0.07007 |
| rs711830 | 2 | 177037311 | G | A | 0.319 | 0.1062 |
| rs62274041 | 3 | 156435640 | A | G | 0.95183 | -0.3712 |
| rs10069690 | 5 | 1279790 | C | T | 0.2598 | 0.08304 |
| rs78724141 | 8 | 82659661 | G | T | 0.06752 | 0.1518 |
| rs10088218 | 8 | 129543949 | G | A | 0.132 | -0.1622 |
| rs7032221 | 9 | 16914895 | T | C | 0.3212 | -0.1773 |
| rs635634 | 9 | 136155000 | C | T | 0.1972 | 0.09884 |
| rs1802669 | 10 | 21827796 | G | A | 0.3482 | 0.0823 |
| rs7135337 | 12 | 121404155 | A | C | 0.5822 | -0.05973 |
| rs11651755 | 17 | 36099840 | T | C | 0.4865 | 0.03128 |
| rs1105569 | 17 | 43793388 | C | T | 0.2185 | 0.113 |
| rs7207826 | 17 | 46500673 | T | C | 0.2678 | 0.1048 |
| rs61494113 | 19 | 17401859 | C | T | 0.2976 | 0.1198 |
| rs9625477 | 22 | 28858248 | T | C | 0.09695 | -0.1045 |

*Table s5. List of the 15 common variants genotyped in the UK Collaborative Trial of Ovarian Cancer Screening (UKCTOCS) that were used to construct a PRS for model validation.*

### Results

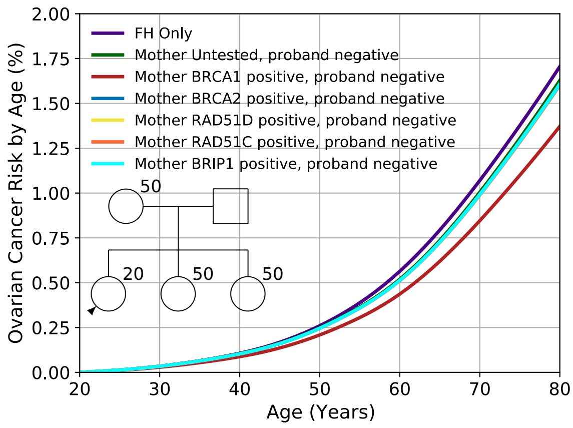


(a)

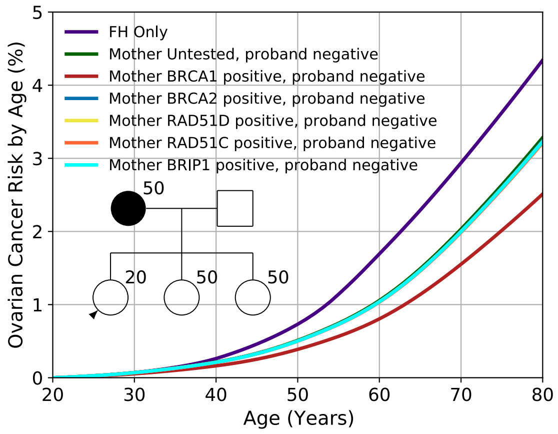


(b)

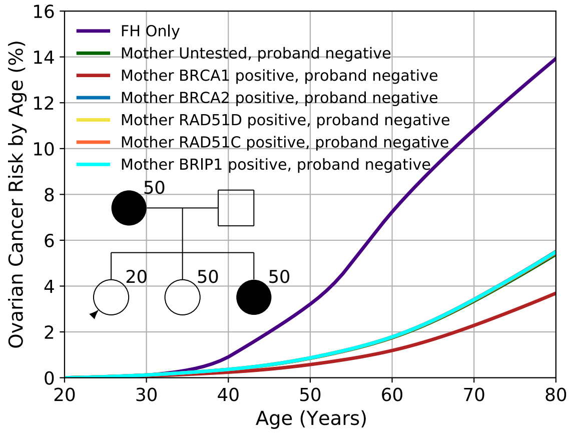


(c)

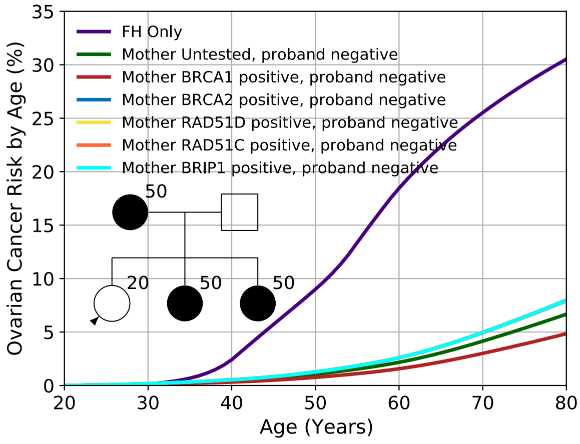


(d)

*Figure s1. The effect of negative-predictive testing in the proband. Predicted lifetime (age 20 to 80 years) EOC risk by family history and PV in the mother and proband. Each Figure ((a)-(d)) shows the risk based on the PV status of the mother assuming the proband tests negative for the PV the mother carries. In the case of "untested" mother, the proband is assumed to test negative for all genes. In the case of "FH Only", neither the proband nor her mother are tested. Screening test sensitivities are set to 1.0 for all genes. The corresponding values are given in Table s6.*

| SCREENING RESULT\FH | (A) | (B) | (C) | (D) |
|---|---|---|---|---|
| **FH ONLY** | 1.7 | 4.3 | 13.9 | 30.5 |
| **UNTESTED** | 1.6 | 3.3 | 5.4 | 6.7 |
| ***BRCA1* PV** | 1.4 | 2.5 | 3.7 | 4.8 |
| ***BRCA2* PV** | 1.6 | 3.2 | 5.5 | 8.0 |
| ***RAD51D* PV** | 1.6 | 3.2 | 5.5 | 8.0 |
| ***RAD51C* PV** | 1.6 | 3.2 | 5.5 | 7.9 |
| ***BRIP2* PV** | 1.6 | 3.2 | 5.5 | 7.9 |

*Table s6. Predicted lifetime ovarian cancer risk for the different family histories (FH) shown in Figure s1 based on the screening test results for pathogenic variants (PVs) in BRCA1, BRCA2, RAD51D, RAD51C and BRIP1, where the proband tests negative for PVs in all genes, and her mother tests positive for a PV in the indicated gene. In the case of "untested", the proband tests negative for all genes, while the mother is untested. In the case of "FH Only", neither the proband nor her mother are tested. Each Figure is based on the same family structure, but with an increasing number of affected first-degree relatives, as indicated by the insert pedigree diagrams in Figure s1.*

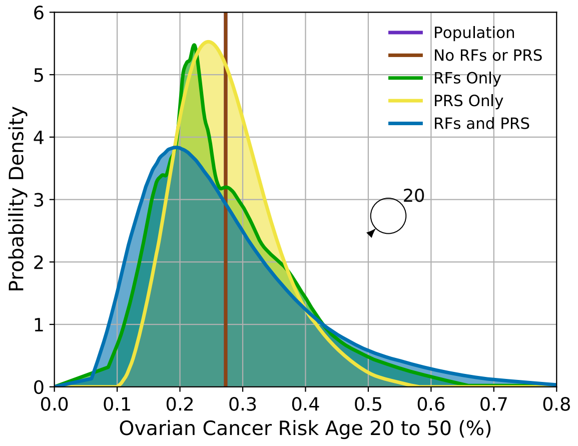


**(a)**

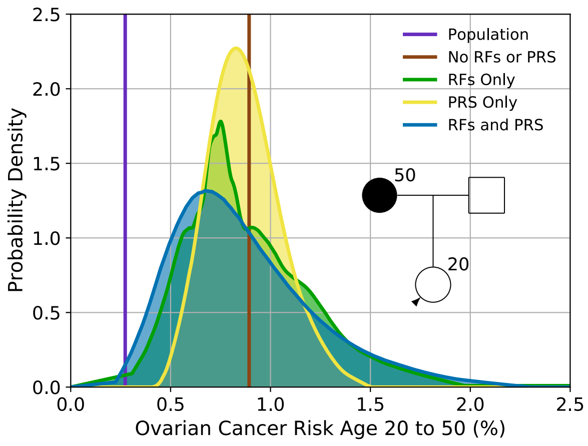


**(b)**

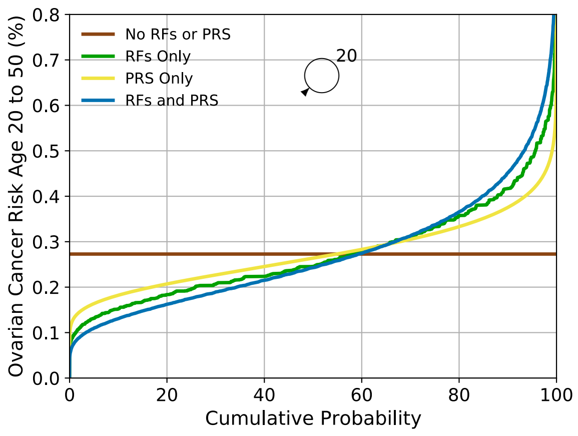


**(c)**

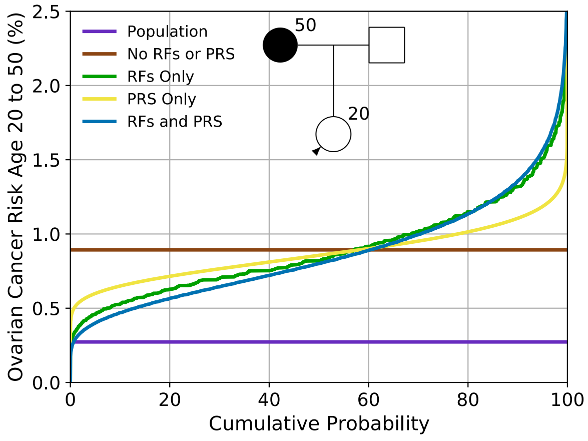


**(d)**

*Figure s2. Predicted EOC risk by age 50 for a female untested for pathogenic variants based on the different predictors of risk (epidemiological risk factors (RFs) and PRS). Figures (a) and (c) show the risk for a female with an unknown family history (equivalent to the distribution of risk in the population), while Figures (b) and (d) show the risk for a female with a mother affected at age 50. Figures (a) and (b) show the probability density function against absolute risk, while Figures (c) and (d) show absolute risk against cumulative distribution. The vertical line in Figure (a) and the horizontal line in Figure (c) (labelled "no RFs or PRS") is equivalent to the population risk of EOC. The "Population" risk is shown separately in Figures (b) and (d). Predictions are based on UK EOC population incidences.*

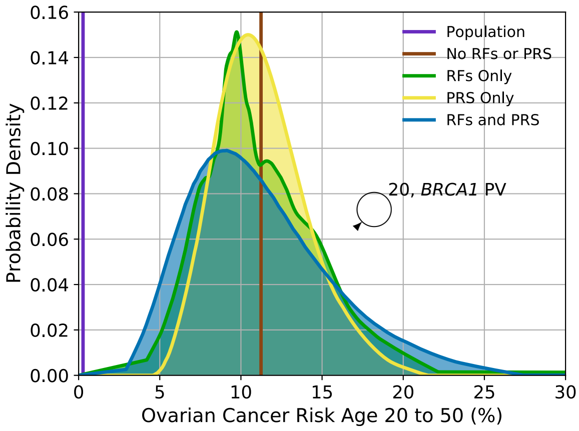
Population
No RFs or PRS
RFs Only
PRS Only
RFs and PRS
20, *BRCA1* PV
Probability Density
Ovarian Cancer Risk Age 20 to 50 (%)

(a)

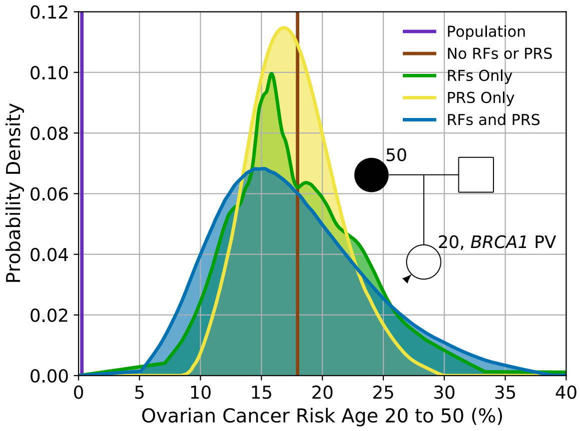
Population
No RFs or PRS
RFs Only
PRS Only
RFs and PRS
50
20, *BRCA1* PV
Probability Density
Ovarian Cancer Risk Age 20 to 50 (%)

(b)

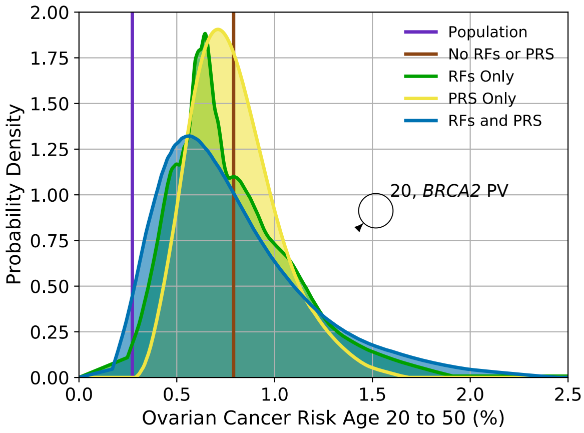
Population
No RFs or PRS
RFs Only
PRS Only
RFs and PRS
20, *BRCA2* PV
Probability Density
Ovarian Cancer Risk Age 20 to 50 (%)

(c)

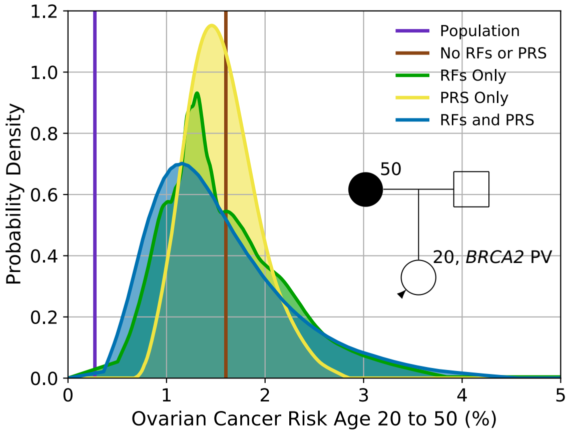
Population
No RFs or PRS
RFs Only
PRS Only
RFs and PRS
50
20, *BRCA2* PV
Probability Density
Ovarian Cancer Risk Age 20 to 50 (%)

(d)

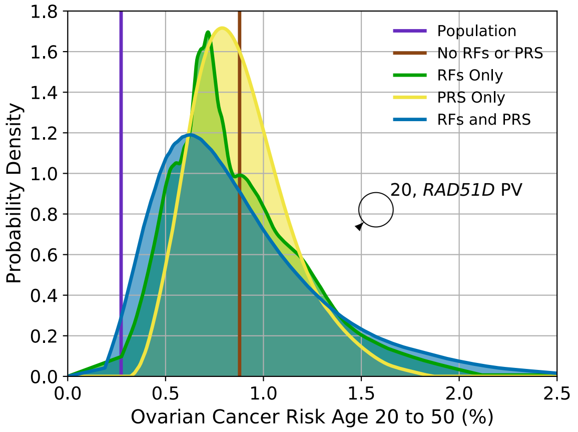
Population
No RFs or PRS
RFs Only
PRS Only
RFs and PRS
20, *RAD51D* PV
Probability Density
Ovarian Cancer Risk Age 20 to 50 (%)

(e)

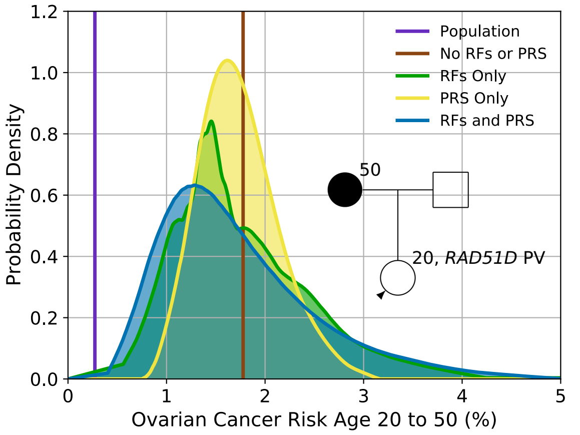
Population
No RFs or PRS
RFs Only
PRS Only
RFs and PRS
50
20, *RAD51D* PV
Probability Density
Ovarian Cancer Risk Age 20 to 50 (%)

(f)

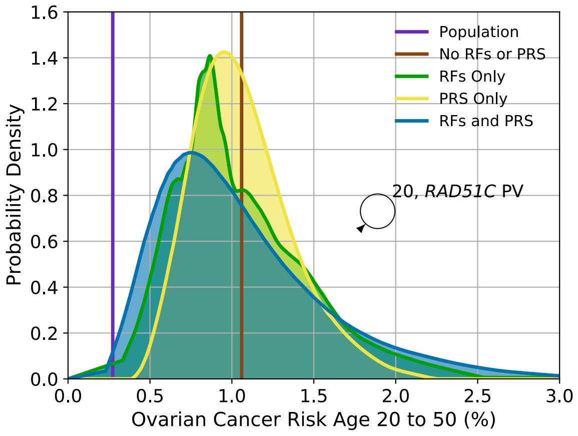


**(g)**

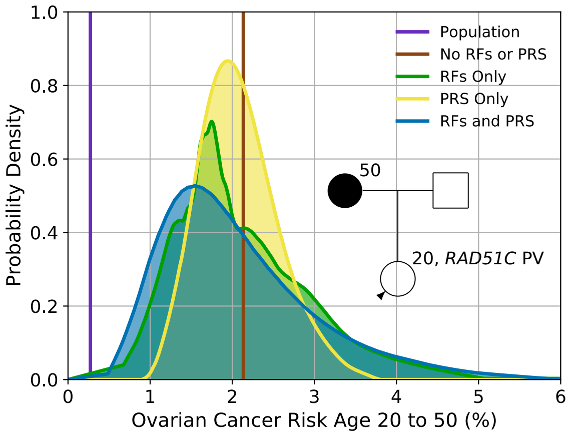


**(h)**

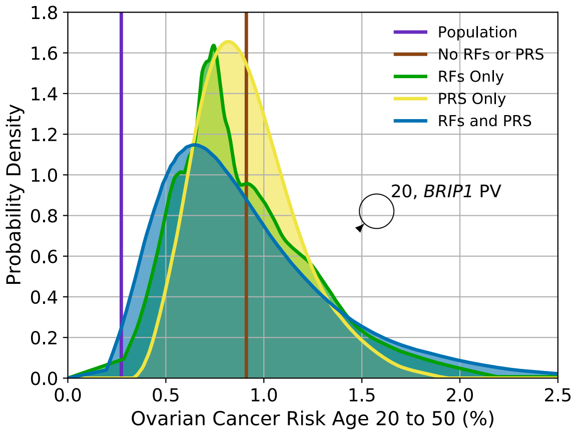


**(i)**

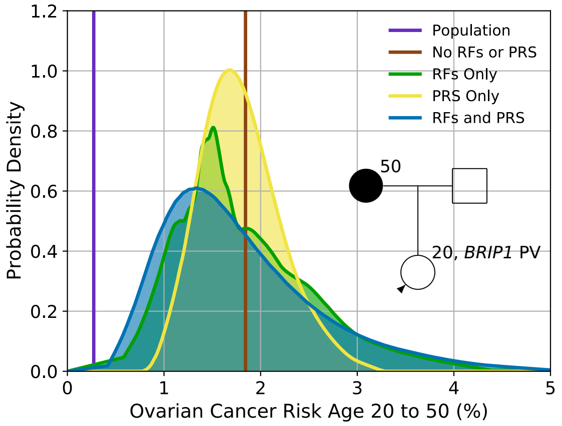


**(j)**

*Figure s3. Predicted ovarian cancer risk by age 50 for a female who has a pathogenic variant (PV) in one of the high or intermediate risk genes included in the model, based on the different predictors of risk (epidemiological risk factors (RFs) and PRS), for two family histories. Figures (a) and (b) show the for a carrier of a PV in* BRCA1*, Figures (c) and (d) show the risk for a carrier of a PV in* BRCA2*, Figures (e) and (f) show the risk for a carrier of a PV in* RAD51D*, Figures (g) and (h) show the risk for a carrier of a PV in* RAD51C*, while Figure (i) and (j) show the risk for a carrier of a PV in* BRIP1*. Figures (a), (c), (e), (g) and (i) show risks for an unknown family history, while Figures (b), (d), (f), (h) and (j) show risks for a female whose mother diagnosed with EOC at age 50. Predictions based on UK ovarian cancer incidences.*

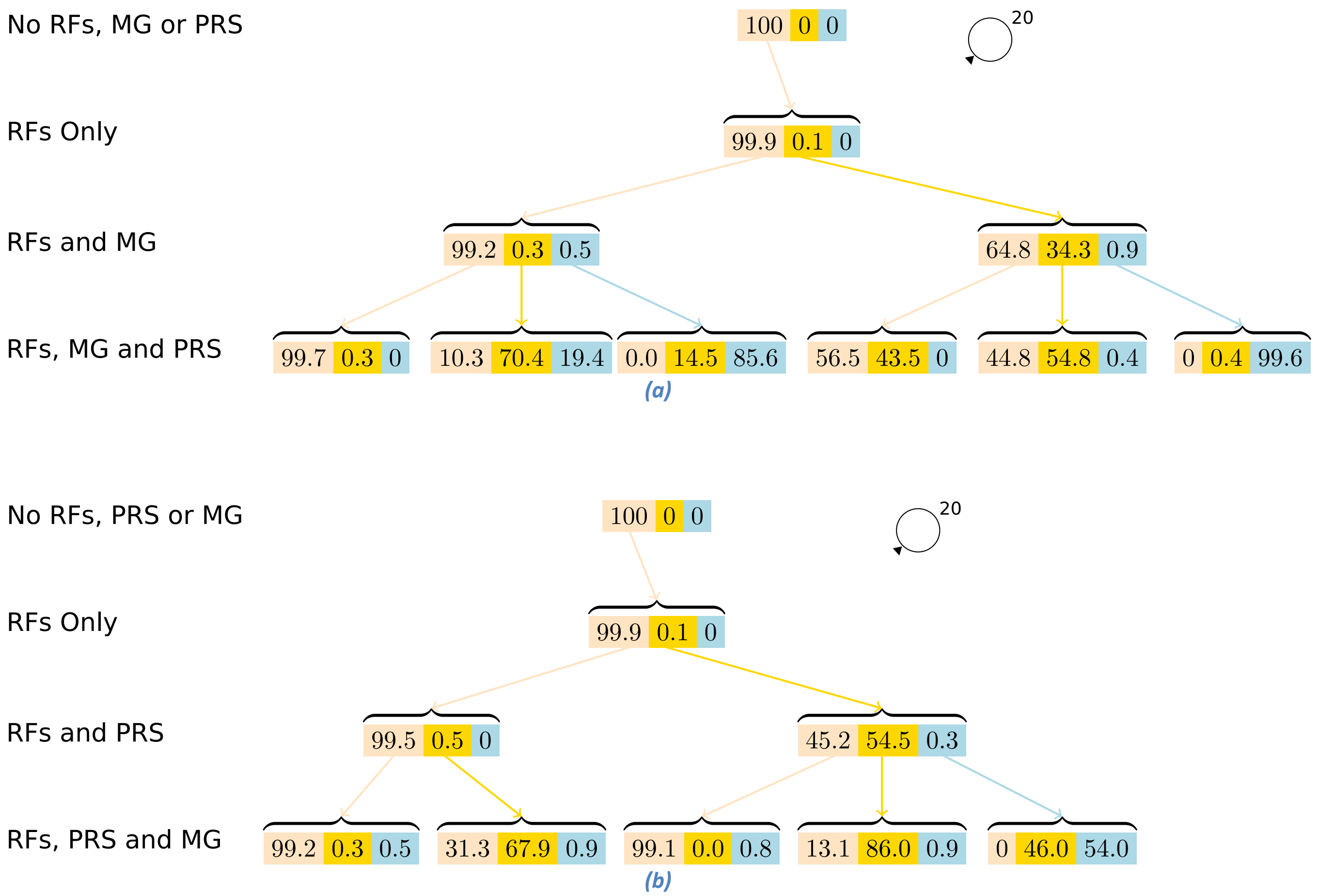


*Figure s4. Probability Trees for lifetime risks, for a female with unknown family history. Starting at the top of each tree, the Figures show the per cent of females reclassified by adding in more information to the EOC risk prediction, as indicated by the captions on the left (epidemiological risk factors (RFs), Polygenic Risk Score (PRS) and testing for pathogenic variants in the major genes (MG)). In Figure (a) MG is added before PRS, while in Figure (b) the PRS is added before MG. Each triplet of numbers is the percentage of females who fall into the risk categories: 1) near population risk shaded in pink ($< 5\%$), 2) moderate risk shaded in yellow ($\geq 5\%$ and $< 10\%$) and 3) high risk, shaded in blue ($\geq 10\%$). "0" percentages indicate that no females fall into that category), while percentages less than 0.1 are denoted by "0.0" (i.e., a very small number of females fall into that category).*

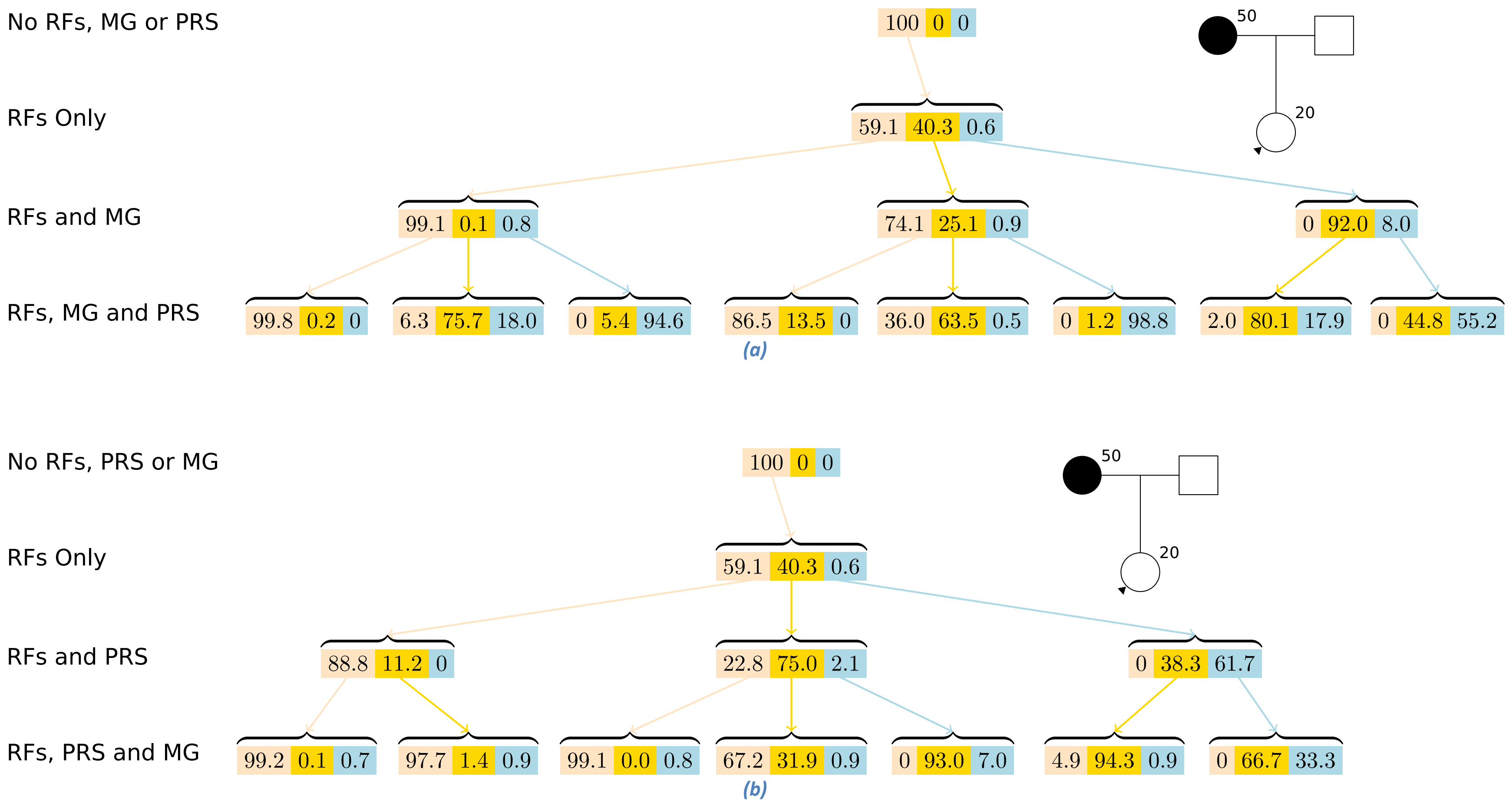


*Figure s5. Probability Trees for lifetime risks, for a female with a mother diagnosed with EOC at age 50. Starting at the top of each tree, the Figures show the per cent of females reclassified by adding in more information to the EOC risk prediction, as indicated by the captions on the left (epidemiological risk factors (RFs), Polygenic Risk Score (PRS) and testing for pathogenic variants in the major genes (MG)). In Figure (a) MG is added before PRS, while in Figure (b) the PRS is added before MG. Each triplet of numbers is the percentage of females who fall into the risk categories: 1) near population risk shaded in pink (<5%), 2) moderate risk shaded in yellow (≥5% and <10%) and 3) high risk, shaded in blue (≥10%). "0" percentages indicate that no females fall into that category), while percentages less than 0.1 are denoted by "0.0" (i.e., a very small number of females fall into that category).*

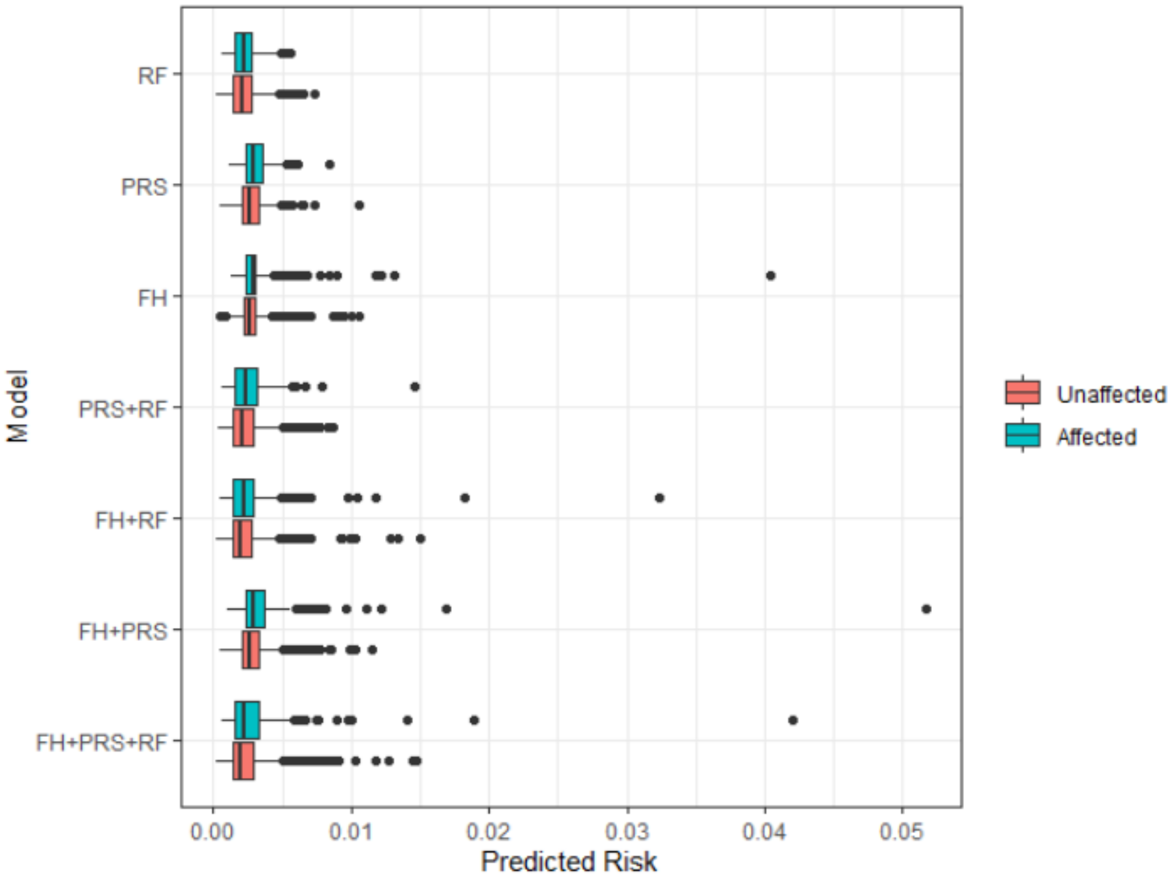


*Figure s6. Predicted 5-year risk distributions considering different information. (RF: epidemiological risk factors; PRS: polygenic risk score; FH: family history).*
